# Automated Patient Identity Verification Framework for Multimodal Medical Imaging Using Deep Metric Learning and Domain Adaptation

**DOI:** 10.64898/2026.08.11.26360177

**Authors:** Yasuyuki Ueda, Takayuki Ishida

## Abstract

**Purpose:** Patient identity management is fundamental in healthcare information systems, as identification inconsistencies can compromise patient safety, data integrity, and clinical workflow efficiency. Reliable linkage of medical images acquired across different imaging modalities remains challenging because of variations in image appearance, acquisition geometry, and imaging characteristics. In this study, we developed an automated patient identity verification framework for multimodal medical imaging using deep metric learning and data-augmented domain adaptation (DADA).

**Methods:** The proposed framework learned modality-invariant patient representations from labeled source-domain data while leveraging unlabeled target-domain data to mitigate cross-modality distribution shifts. Although patient identity annotations were available within the research dataset for evaluation purposes, target-domain labels were intentionally excluded from the DADA optimization process. Chest radiographs and computed tomography (CT) scout images obtained under routine clinical conditions were retrospectively collected and used for evaluation. Verification performance was assessed using receiver operating characteristic (ROC) analysis, with the area under the ROC curve (AUC) used as the primary performance metric.

**Results:** The proposed framework achieved consistently high verification performance across all evaluation conditions, with AUC values ranging from 0.9989 to 0.9999. Similarity-score distributions demonstrated distinct separation between same-patient and different-patient image pairs despite substantial differences between imaging modalities.

**Conclusion:** These findings indicate that patient-specific anatomical representations can be preserved across heterogeneous imaging domains through metric learning and domain adaptation. The proposed framework may serve as a practical infrastructure component for patient identity management, multimodal data integration, quality assurance, and patient safety applications within healthcare information systems.

## 1 Introduction

Accurate patient identification is fundamental in modern healthcare information systems (HISs). Contemporary HIS infrastructures, including electronic health records (EHRs), picture archiving and communication systems (PACSs), vendor-neutral archives, and regional information-sharing networks, depend on reliable patient identity management to ensure data integrity, continuity of care, and safe clinical practice [1–3]. However, the increasing digitization of healthcare has expanded the volume, complexity, and diversity of patient information, making accurate linkage of healthcare data an essential requirement for both clinical care and secondary use of medical data [1–2].

Inaccurate patient matching or information linkage can compromise patient safety, reduce clinical workflow efficiency, and adversely affect healthcare quality [4, 5]. Health information exchange initiatives and interoperable healthcare architectures have enabled the integration of healthcare information across institutions and information systems [6–8]. The growing scale of healthcare data repositories has further increased the need of reliable data quality management and patient-level data integration [9, 10]. Consequently, robust approaches for validating patient identity across heterogeneous healthcare data sources have become increasingly important.

Medical imaging represents one of the largest and most information-rich components of modern healthcare information systems. Large numbers of radiographic, computed tomography (CT), magnetic resonance imaging (MRI), and other diagnostic images are continuously accumulated within institutional archives and exchanged across healthcare networks. Because medical images inherently contain patient-specific anatomical characteristics, image-based patient verification has attracted attention as a complementary approach for patient identity management and quality assurance [11–13].

The use of anatomical information as a biological fingerprint for patient identification has been investigated for several decades. Early studies demonstrated that skeletal structures visible on radiographic examinations could support human identification and patient recognition [14–16]. Subsequent investigations showed the feasibility of preventing filing errors and detecting patient mismatches through automated comparison of chest radiographs stored in PACS environments [17–20]. Similar approaches have also been applied to CT images and other imaging modalities, further supporting the concept that routine medical images contain persistent patient-specific anatomical information suitable for identity verification [21–25].

Recent advances in artificial intelligence (AI) and deep learning have transformed biomedical image analysis, achieving state-of-the-art performance across a wide range of tasks including disease detection, prognostic prediction, image segmentation, and patient representation learning [26, 27]. Deep metric learning (DML) approaches have been successfully applied to chest radiographs, CT images, and other medical imaging modalities, enabling the extraction of discriminative patient representations from routine clinical images [28–35]. These studies suggest that patient-specific anatomical characteristics can be effectively learned by neural networks and used for automated verification.

Despite these advances, most previous studies have focused on patient verification or re-identification within a single imaging modality. In routine clinical practice, however, patients frequently undergo examinations with different imaging modalities. Linking heterogeneous images acquired across different modalities could support patient identity management, multimodal data integration, quality assurance, and patient safety applications within healthcare information systems. Nevertheless, reliable cross-modality patient verification remains challenging because differences in image appearance, acquisition geometry, patient positioning, image contrast, and imaging characteristics introduce significant distribution shifts across imaging modalities.

Domain adaptation (DA) techniques have emerged as an effective strategy for addressing distribution shifts between heterogeneous medical imaging domains [36–38]. By reducing discrepancies between source and target domains, DA enhances representation stability and consistency across disparate domains when labeled target-domain data are limited or unavailable [37, 38]. Recently, data-augmented domain adaptation (DADA) has been proposed as an approach for imparting robustness to proxy-based DML through latent-space augmentation and cross-domain distribution alignment [39]. Because DADA leverages labeled source-domain data together with unlabeled target-domain data, it is particularly attractive for real-world healthcare information systems in which comprehensive patient identity annotations are often unavailable across all imaging modalities.

Furthermore, recent advances in optimization algorithms and neural network architectures have enhanced cross-modality representation learning and improved model generalizability [40, 41]. In particular, DML provides the primary foundation for discriminative identity verification by establishing tight patient-specific clusters in the embedding space [28–35]. Meanwhile, incorporating domain alignment mechanisms—such as maximum mean discrepancy (MMD) employed within DADA [42]—acts as a structural regularizer that mitigates feature distortion caused by domain discrepancies. Furthermore, explainability approaches such as Grad-CAM facilitate interpretation of the anatomical regions used by deep-learning models for verification decisions [43].

Although patient verification has been investigated in chest radiography, CT, MRI, and other imaging modalities [15, 16, 21–25, 28–35], automated patient identity verification between chest radiographs and CT scout images remains largely unexplored. Therefore, this study developed an automated patient identity verification framework for multimodal medical imaging using DML and DADA. The proposed framework combines AdaCos-based metric learning for discriminative anatomical representation with DADA for domain-invariant regularization, aiming to safeguard verification stability against severe distribution shifts between chest radiographs and CT scout images. We hypothesized that while DML primarily drives high identification capability, the complementary integration of DA would impart essential robustness to cross-modality domain gaps without requiring patient identity annotations in the target modality, thereby supporting scalable deployment in healthcare information systems.

## 2 Methods

### 2.1 Overview

An automated patient identity verification framework was developed for cross-modality verification between chest radiographs and CT scout images. The framework integrated DML and DADA to learn patient-specific image representations that are inherently robust to severe modality-related distribution shifts. The overall framework consisted of four components: image acquisition, image preprocessing, representation learning, and patient identity verification.

During model development, both the chest radiographs and CT scout images were used with identity annotations. However, consistent with the DADA framework, patient identity annotations from the target modality were not used during DA. The target-domain images were incorporated solely as unlabeled samples for feature-distribution alignment. The trained model generates modality-invariant patient embeddings that can be compared using similarity analysis for automated patient verification. The overall framework is illustrated in **Fig. 1** for model development and in **Fig. 2** for the verification workflow.

**Fig. 1.**
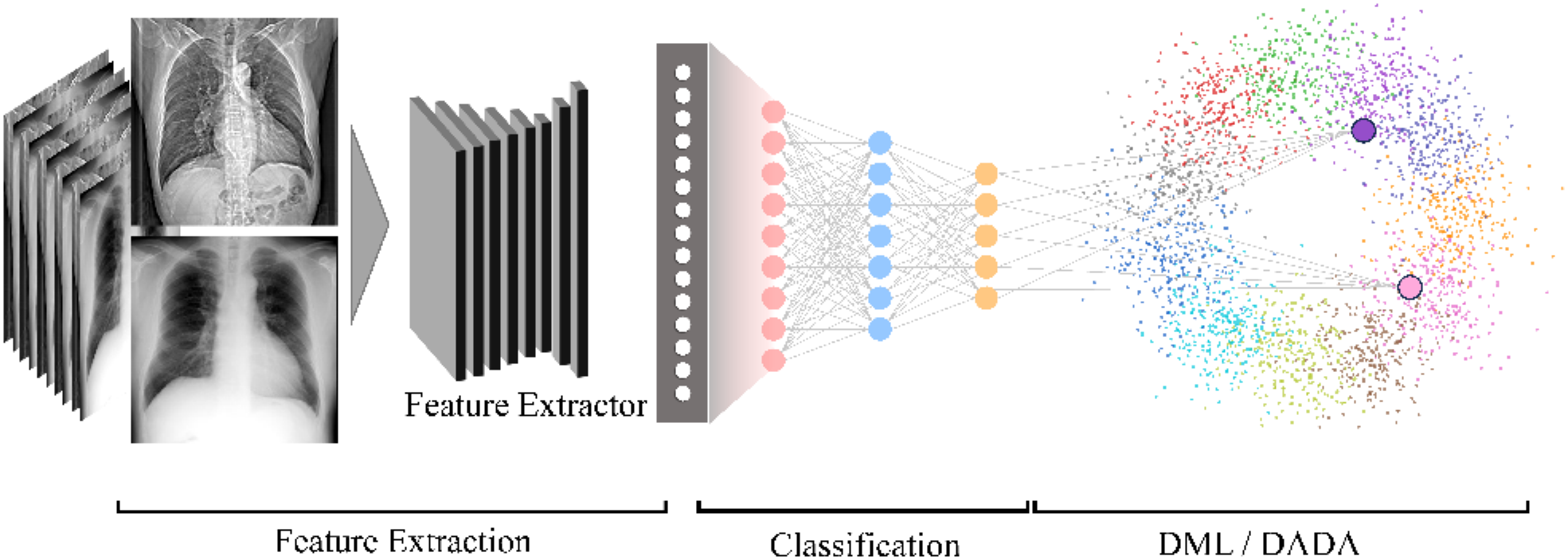
Overview of the proposed framework with DML and DADA for cross-modality patient identity verification, illustrating the image acquisition, preprocessing, representation learning, domain adaptation, and verification processes. Chest radiographs are first processed using a feature extractor to generate the latent feature representations. These features are subsequently used for classification and further optimized via DML and DADA learning. The learned embedding space groups images from the same class closer together, while increasing the separation between the different classes, resulting in more discriminative feature representations. **DML**: Deep Metric Learning; **DADA**: Data-augmented Domain Adaptation

**Fig. 2.**
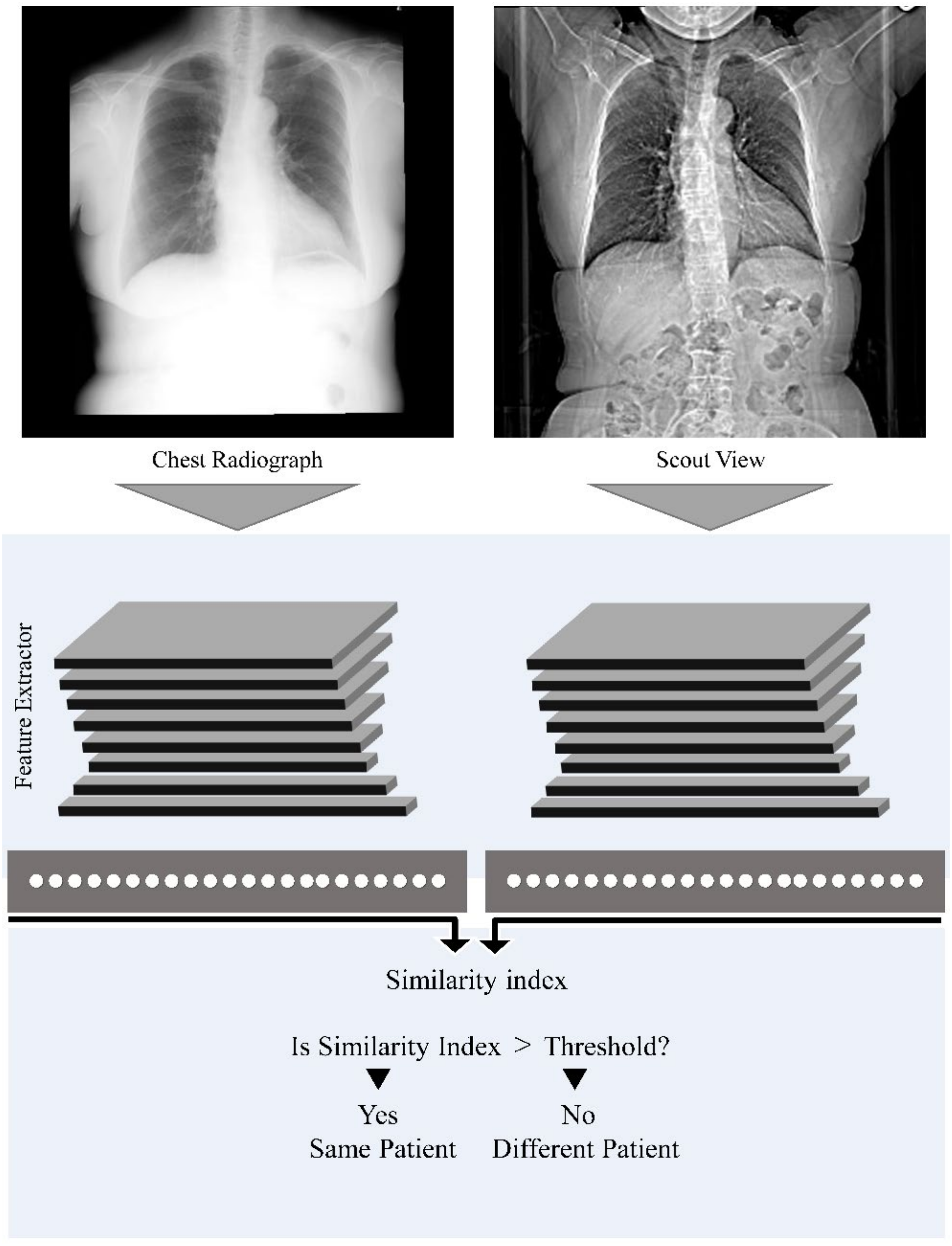
Overview of the proposed method for patient verification using chest radiographs and trunk CT scout images. Input chest radiograph and CT scout images are first transformed into feature embeddings using a trained feature extractor. The similarity score between the two feature embeddings is subsequently computed and compared with a predefined threshold. If the similarity score is equal to or greater than the threshold, the image pair is classified as originating from the same patient; otherwise, it is classified as originating from different patients

### 2.2 Dataset

A retrospective multimodal imaging dataset was used for framework development and evaluation. The dataset comprised chest radiographs and CT scout images acquired from patients who underwent routine chest radiography and trunk CT examinations at Yamaguchi University Hospital, Japan between March 2015 and March 2017 (inclusive).

For each patient, the dataset included chest radiographs acquired under routine clinical follow-up conditions. The dataset also included trunk CT scout images acquired using three clinical CT scanners from two vendors.

This study used de-identified images retrospectively retrieved from the PACS of Yamaguchi University Hospital. All research activities, including study design, algorithm development, and data analysis, were conducted at the University of Osaka. This study was approved by the Institutional Review Board of the University of Osaka Hospital (Approval No. 21064–6). The requirement for informed consent was waived owing to the retrospective nature of the study.

To ensure complete independence between the training/validation and final testing processes, the dataset was split at the patient level. Patients with one or two examinations were assigned exclusively to the independent holdout test set (11,589 patients; 14,469 images), while patients with three or more examinations (3,203 patients; 19,885 images) were allocated to the development cohorts for five-fold cross-validation. In each fold, the development cohort was partitioned into training and validation subsets at an 80:20 patient ratio in a completely patient-disjoint manner (2,563 patients in training and 640 patients in validation per fold; **Table 1**). Consequently, there is zero patient overlap not only between the development and test sets but also between the training and validation subsets within each fold.

**Table 1.** Patient demographics of datasets used for training, validation, and testing.

| Fold | Subset | Number of images | Number of patients | Age, years |  | Sex |  |
|---|---|---|---|---|---|---|---|
|  |  |  |  | mean | SD | male | female |
| **Total** |  | 34,354 | 14,792 | 64.5 | 15.8 | 7,821 | 6,971 |
| 1 | **Training** | 15,985 | 2,563 | 67.2 | 13.8 | 1,532 | 1,031 |
|  | **Validation** | 3,900 | 640 | 67.5 | 13.5 | 389 | 251 |
| 2 | **Training** | 15,785 | 2,563 | 67.2 | 13.8 | 1,540 | 1,023 |
|  | **Validation** | 4,100 | 640 | 67.2 | 13.2 | 381 | 259 |
| 3 | **Training** | 15,915 | 2,563 | 67.2 | 13.6 | 1,552 | 1,011 |
|  | **Validation** | 3,970 | 640 | 67.3 | 14.0 | 369 | 271 |
| 4 | **Training** | 16,009 | 2,563 | 67.4 | 13.6 | 1,522 | 1,041 |
|  | **Validation** | 3,876 | 640 | 66.7 | 14.2 | 399 | 241 |
| 5 | **Training** | 15,846 | 2,560 | 67.2 | 13.7 | 1,538 | 1,022 |
|  | **Validation** | 4,039 | 643 | 67.5 | 13.6 | 383 | 260 |
| **Independent Test** |  | 14,469 | 11,589 | 63.8 | 16.2 | 5,900 | 5,689 |

**Note:** Patients who underwent three or more examinations (*n* = 3,203; 19,885 images) were included in the five-fold cross-validation development cohort and partitioned into training and validation sets at an 80:20 patient ratio ( 2,563 patients in training and 641 patients in validation per fold) in a patient-disjoint manner. Patients who underwent one or two examinations (*n* = 11,589; 14,469 images) constituted the independent holdout test cohort. \*Values for five-fold training and validation sets are presented as mean and SD across the five folds.
**SD**: standard deviation.

Model training was conducted on individual images using a proxy-based metric learning framework (AdaCos) without any explicit pair generation. For each patient in the test set, the earliest available examination was designated as the baseline image, with subsequent examinations treated as follow-up images. For model evaluation, positive pairs comprised images from the same patient, while negative pairs comprised images from different patients, yielding 2,880 same-patient pairs and 33,373,440 theoretically possible different-patient pairs in the test set. Patient demographics and imaging conditions are summarized in **Tables 1** and **2**.

**Table 2.** Imaging parameters and acquisition conditions for chest radiography and CT scout imaging in the dataset.

| **Imaging method** | **Scout view** |  | **Radiography** |  |
|---|---|---|---|---|
| **Examined region** | Trunk, including the chest |  | Chest |  |
| **Vendor** | SMS | GEMS | Konica Minolta Inc. |  |
| **View position** | PA |  | AP | PA |
| **Number of patients** | 4,762 | 1,553 | 619 | 10,380 |
| **Number of images** | 7,073 | 1,778 | 1,589 | 23,914 |
| **Pixel spacing (mm), (columns × rows)** | 2.0 × 2.0 | 0.55 × 0.60 | 0.175 × 0.175 |  |

**Note: SMS**: Siemens Medical Solutions; **GEMS**: GE Medical Systems; **PA**: posteroanterior; **AP**: anteroposterior.

### 2.3 Deep Metric Learning Framework

The patient identity verification task was formulated as a DML problem. The objective of the framework was to learn an embedding space in which images belonging to the same patient are positioned close to one another, whereas images from different patients are separated. To enhance cross-modality representation robustness and prevent feature distortion caused by domain discrepancy, DADA was incorporated as a domain alignment regularizer during model training. DADA aligns feature distributions across source, target, and augmented domains, thereby reducing the domain discrepancy between chest radiographs and CT scout images. To isolate and evaluate the regularizing contribution and domain-invariant robustness imparted by DADA, baseline metric learning without DA (AdaCos alone) and standard maximum mean discrepancy (MMD) alignment were also investigated as ablation models. Detailed mathematical descriptions of feature embedding, DA, MMD, and optimization objectives are provided in Online Resource 1 (**Supplementary Methods S1**).

### 2.4 Network Implementation Details

#### 2.4.1 Feature Extractor

EfficientNetV2-S was adopted as the backbone feature extractor because of its favorable balance between representational capacity and computational efficiency [41]. The network was trained from scratch without external pretraining. Feature maps from the final convolutional block were transformed into embedding vectors and subsequently used for metric-learning optimization and similarity analysis.

#### 2.4.2 Data Augmentation

Data augmentation techniques—including random rotation, transversal scaling of CT scout images, and perspective distortion of chest radiographs—were applied during training to improve the generalization performance and reduce overfitting. Detailed augmentation procedures are provided in Online Resource 1 (Supplementary Methods S2).

#### 2.4.3 Domain Adaptation Strategy

Consistent with the original DADA framework [39], feature distributions from source, target, and augmented domains were aligned through MMD-based optimization. This strategy penalizes distribution discrepancies in the latent space, functioning as a structural constraint that stabilizes cross-modality patient embeddings against appearance variations and scanner-specific differences.

#### 2.4.4 Training Configuration

The model was trained using Adaptive Sharpness-Aware Minimization (ASAM) [40] for up to 500 epochs without any early stopping. Training was conducted with a mini-batch size of 16 (effective batch size of 64 via gradient accumulation) and an initial learning rate of 0.01, which decayed to 0.0001 following a cosine schedule. For the primary evaluation of the proposed framework, training was conducted independently across all five folds. In each fold, open-set patient re-identification performance was evaluated on the patient-disjoint validation cohort at every epoch, and the checkpoint achieving the highest validation top-1 accuracy (ACC@1) was retained for final evaluation on the independent holdout test set. Performance metrics across the five folds are reported as the mean and standard deviation. For the ablation experiments, all comparative model variants were trained on a fixed representative benchmark partition (Fold 1) and evaluated on the identical independent holdout test set to ensure a strictly controlled, paired comparison devoid of data-partitioning variability while maintaining computational feasibility. Additional training hyperparameters are provided in Online Resource 1 (Supplementary Methods S3).

### 2.5 Evaluation Metrics

The performance of the proposed DML framework was evaluated under two operational scenarios: 1:1 matching for patient verification and 1:N matching for open-set patient re-identification. In both scenarios, pairwise similarity between two patient embeddings was computed using cosine similarity:

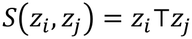

where *z_i_* and *z_j_* denote L2-normalized patient embeddings.

#### 2.5.1 Patient Verification (1:1 Matching)

Patient verification was evaluated across all possible image pairs in the independent test cohort as the primary 1:1 matching endpoint. For each evaluation condition, positive image pairs (*N_s_* = 2,880) consisted of images from the same patient, whereas negative image pairs (*N_i_* = 33,373,440) consisted of images from different patients. To ensure an unbiased and comprehensive assessment without statistical artifacts from down-sampling, exhaustive evaluation across all 33,373,440 negative pairs was conducted. Verification performance was evaluated using receiver operating characteristic (ROC) analysis and the corresponding area under the curve (AUC). In accordance with international biometric performance reporting standards (ISO/IEC 19795-1), we also computed the Equal Error Rate (EER), where the False Non-Match Rate (FNMR) equals the False Match Rate (FMR), and verification sensitivities (True Match Rates: 1 − FNMR) at operationally stringent decision thresholds corresponding to FMRs of 1.0% (@FMR100) and 0.1% (@FMR1000).

#### 2.5.2 Patient Re-Identification (1:N Matching)

To assess practical utility in clinical image retrieval and misfiling detection, open-set 1:N patient re-identification was evaluated. In this setting, each follow-up image was matched against a reference database consisting of one baseline examination per patient. A follow-up image was deemed correctly re-identified if the true corresponding patient yielded the highest cosine similarity among all candidate patients (Top-1 accuracy (ACC@1) on the cumulative match characteristic curve).

#### 2.5.3 Anatomical Masking Analysis

To confirm that the proposed framework relied on genuine thoracic anatomical biometrics rather than non-anatomical shortcut learning (such as peripheral collimator borders, scanner-specific edge characteristics, lead orientation markers, or burned-in DICOM text annotations), an anatomical masking experiment was conducted on the independent test cohort. The squared lung-field bounding box was automatically delineated using a pre-trained PSPNet segmentation model from TorchXRayVision [44]. Models trained with the default top-centered full field of view (Full FoV) were compared against those trained and evaluated strictly on the cropped lung-field region of interest (Lung-field ROI), wherein all outer peripheral margins and device artifacts outside the thoracic cage were excluded.

### 2.6 Statistical Analysis

For the primary evaluation using five-fold cross-validation, performance metrics are reported as the mean ± standard deviation across the five trained models. For the ablation study, differences in ACC@1 on paired test queries were evaluated using McNemar’s test (two-sided, with statistical significance set at p < 0.05). All statistical analyses and metric computations were conducted using Python (version 3.12.11) with scikit-learn (version 1.4.0) and SciPy (version 1.12.0).

## 3 Results

The performance of the proposed framework was evaluated on the independent test cohort (11,589 patients; 14,469 images) using five-fold cross-validation models. For each fold, the model checkpoint achieving the highest validation ACC@1 was selected. Primary performance metrics across the five folds and ablation comparisons are summarized in **Tables 3** and **4**, respectively.

**Table 3.** Patient identity verification and re-identification performance of the proposed framework evaluated across imaging modalities and intrinsic domain conditions on the independent test dataset.

| D1 | D2 | *N<sub>s</sub>* | ACC@1 (%) | AUC | EER (%) | Sensitivity (%) |  |
|---|---|---|---|---|---|---|---|
|  |  |  |  |  |  | @FMR100 | @FMR1000 |
| All | — | 2,880 | 98.3 ± 0.131 | 0.9998 ± 0.0000 | 0.359 ± 0.042 | 99.8 ± 0.067 | 99.2 ± 0.093 |
| Cross-modality | — | 334 | 93.6 ± 0.781 | 0.9999 ± 0.0000 | 0.423 ± 0.160 | 99.9 ± 0.164 | 99.0 ± 0.164 |
| Same-modality | All | 2,546 | 98.9 ± 0.133 | 0.9998 ± 0.0001 | 0.416 ± 0.066 | 99.8 ± 0.076 | 99.2 ± 0.083 |
| Chest Radiography | Same View-position | 1,518 | 99.8 ± 0.086 | 0.9999 ± 0.0001 | 0.171 ± 0.059 | 99.9 ± 0.055 | 99.8 ± 0.059 |
|  | Different View-position | 40 | 81.5 ± 2.236 | 0.99974 ± 0.0001 | 0.222 ± 0.135 | 100.0 ± 0.000 | 93.5 ± 2.236 |
| Scout View | Same-Vendor | 651 | 98.7 ± 0.296 | 0.9999 ± 0.0000 | 0.434 ± 0.071 | 99.9 ± 0.137 | 99.1 ± 0.188 |
|  | Cross-Vendor | 337 | 96.9 ± 0.615 | 0.9989 ± 0.0004 | 0.653 ± 0.133 | 99.5 ± 0.133 | 98.9 ± 0.163 |

**Note: D1**: imaging modality; **D2:** domain intrinsic to the modality (view position for chest radiography and scanner vendor for CT scout views); ***N<sub>s</sub>***: number of same-patient pairs; **ACC@1**: top-1 accuracy; **SMS**: Siemens Medical Solutions; **GEMS**: GE Medical Systems; PA: posteroanterior; **AP**: anteroposterior; **AUC**: area under the receiver operating characteristic curve; **EER**: equal error rate; **Sensitivity:** true match rate (*1 - FNMR*); **@FMR100**: at a FMR of 1.0%; **@FMR1000**: at a FMR of 0.1%; **FNMR**: false non-match rate; **FMR**: false match rate.

**Table 4.** Performance comparison of ablation models assessing the impact of Data-Augmented Domain Adaptation and lung-field cropping on patient re-identification using the independent test dataset.

| Model / Configuration | Alignment Method | Input FoV | ACC@1(%) |  |  |
|---|---|---|---|---|---|
|  |  |  | Overall<br>(*N<sub>s</sub>* = 2,880) | Same-Modality<br>(*N<sub>s</sub>* = 2,546) | Cross-Modality<br>(*N<sub>s</sub>* = 334) |
| Proposed Framework | DADA (lambda=0.05) | Full FoV | 98.3 | 98.7 | 94.6 |
| (A) AdaCos without DADA | None | Full FoV | 98.3 | 98.9 | 93.7 |
| (B) MMD-based Alignment | MMD (lambda=0.05) | Full FoV | 98.2 | 98.9 | 92.5 |
| (C) Anatomical Crop Model | DADA (lambda=0.05) | Lung-field ROI | 98.2 | 98.8 | 93.1 |

**Note:** Full FoV denotes the default top-centered 448 × 448 mm² field of view covering the upper-to-mid thoracic region without anatomical cropping. Lung-field ROI denotes images cropped specifically around the segmented lung bounding box to exclude peripheral thoracic borders and scanner margins.
**FoV**: field of view; ***N<sub>s</sub>*:** number of same-patient pairs; **ACC@1**: top-1 accuracy; **ROI**: region of interest.

### 3.1 Patient Verification and Re-Identification Performance

Patient verification and re-identification performance on the test cohort are summarized in **Table 3**. Across the five folds, the overall verification performance yielded an AUC of 0.9998 ± 0.0000 and an ACC@1 of 98.3 ± 0.131%. The EER was 0.359 ± 0.042%, with sensitivities of 99.8 ± 0.067% at FMR100 and 99.2 ± 0.093% at FMR1000. Under cross-modality evaluation, the AUC was 0.9999 ± 0.0000 and the ACC@1 was 93.6 ± 0.781%. Under same-modality evaluation, the corresponding values were 0.9998 ± 0.0001 and 98.9 ± 0.133%, respectively. In chest radiography subgroups comparisons, evaluations acquired with the same view position yielded an ACC@1 of 99.8 ± 0.086% and an AUC of 0.9999 ± 0.0001, whereas those involving different view positions (PA vs. AP) yielded an ACC@1 of 81.5 ± 2.24% and an AUC of 0.9998 ± 0.0001. In CT scout images, same-vendor comparisons achieved an ACC@1 of 98.7 ± 0.296% and an AUC of 0.9999 ± 0.0000, whereas cross-vendor comparisons achieved an ACC@1 of 96.9 ± 0.615% and an AUC of 0.9989 ± 0.0004.

### 3.2 Ablation Study of Domain Adaptation Framework

Ablation analysis comparing AdaCos alone (None), AdaCos with MMD, and DADA trained on the representative benchmark partition (Fold-1) is summarized in **Table 4**. Overall ACC@1 was 98.3% for AdaCos alone, 98.2% for MMD, and 98.3% for DADA; the difference between DADA and AdaCos alone did not reach statistical significance (McNemar’s test, p > 0.05). Cross-modality ACC@1 was 93.7% for AdaCos alone, 92.5% for MMD, and 94.6% for DADA.

### 3.3 Grad-CAM Visualization and Anatomical Masking Analysis

Grad-CAM activations were localized to internal thoracic anatomical structures across both chest radiographs and CT scout images (Online Resource 1, **Figure S1**). In the anatomical masking experiment, all peripheral margins outside the segmented lung bounding box were excluded. As presented in **Table 4**, overall ACC@1 was 98.3% with the full FoV and 98.2% with the lung-field ROI. Same-modality ACC@1 was 98.7% with the full FoV and 98.8% with the lung-field ROI. Cross-modality ACC@1 was 94.6% with the full FoV and 93.1% with the lung-field ROI.

## 4 Discussion

### 4.1 Principal Findings

This study demonstrated that patient-discriminative anatomical representations can be learned and transferred across heterogeneous imaging modalities for automated identity verification, even when patient identity annotations are unavailable in the target domain through domain adaptation [36–39]. While prior studies have established the feasibility of image-based patient verification within single modalities [15, 16, 21–25, 28–35], our findings extend this capability across disparate modalities. Notably, the ablation analysis revealed that identification accuracy was largely driven by AdaCos-based metric learning, which pushed overall ACC@1 to near-ceiling levels (>98%). Consequently, the performance difference between DADA and AdaCos alone in cross-modality ACC@1 (94.6% vs. 93.7%) did not reach statistical significance (p > 0.05). Furthermore, standard MMD-based alignment without latent data augmentation yielded a slightly lower cross-modality ACC@1 (92.5%), highlighting that naive distribution alignment without class-aware regularization risks negative transfer by inadvertently compressing inter-patient discrimination margins. In contrast, by synthesizing a manifold bridge between samples and corresponding class proxies via latent mixup, DADA preserves semantic class boundaries during domain adaptation [39,42], thereby preventing feature degradation and achieving the highest cross-modality generalization (94.6%). Rather than serving as an accuracy booster, the primary role of DADA is imparting structural robustness against severe cross-modality distribution shifts [36–39]. While AdaCos maps identity traits into a compact metric space, DADA functions as a latent-space regularizer that aligns cross-modality distributions while preserving class-separability via proxy mix-up, preventing feature distortion arising from differences in acquisition geometry, spatial resolution, and scanner hardware. DADA thus decouples patient biometric signatures from modality-specific artifacts, ensuring representational stability across disparate clinical domains.

### 4.2 Cross-Modality Patient Identity Representation

Cross-modality verification between chest radiographs and CT scout images is inherently challenged by major discrepancies in projection physics, contrast characteristics, and patient positioning [24, 25 ,36–38]. By aligning feature distributions across source, target, and augmented domains via data-augmented three-way MMD optimization [39], DADA provided stable representation learning across different scanner vendors and radiographic view orientations. This divergence highlights the fundamentally distinct operational demands of the two tasks: verification evaluates a pairwise 1:1 match against an established threshold, whereas re-identification requires ranking the correct patient first among an entire 11,589-candidate gallery. The open-set 1:N scenario is therefore substantially more sensitive to minor similarity perturbations across the vast search space.

### 4.3 Implications for Healthcare Information Systems

In modern healthcare information infrastructures (PACS, EHR, and vendor-neutral archives), patient linkage errors across departments and imaging suites present persistent safety and data-integrity risks [1–10]. Because the proposed framework leverages routinely acquired diagnostic images, it requires no dedicated biometric hardware and involves no extra patient burden. The system can function as an automated, background safety gate within PACS workflows to intercept potential misfiling incidents before clinical interpretation, complementing administrative verification procedures [4, 5, 11–13, 17]. Furthermore, reliable cross-modality linkage supports the automated curation of large-scale, multimodal research imaging repositories by verifying identity consistency across longitudinal records [6–10].

### 4.4 Anatomical Interpretability and Shortcut Mitigation

Grad-CAM visualizations confirmed that model attention was predominantly focused on stable upper thoracic skeletal anatomy, particularly the lung apices and clavicular regions, aligning with forensic anthropological literature on radiographic biometrics [14–17,19–23]. Crucially, the anatomical masking experiment (**Table 4**) demonstrated that confining the field of view strictly to the lung-field ROI yielded virtually identical performance to the full FoV. This outcome confirms that the model relies on persistent internal anatomical morphology rather than exploiting non-anatomical shortcuts, such as collimator borders, scanner-specific edge markers, or burned-in annotations.

### 4.5 Limitations

This study has several limitations. First, the dataset was derived from a single academic medical center. Although multiple scanner vendors and imaging protocols were evaluated, external validation across multi-institutional cohorts is necessary to confirm generalizability [21–25, 28–35]. Second, the framework operated on two-dimensional projection images; extending domain-invariant metric learning to full 3D volumetric CT and MRI remains an important research objective. Third, the dataset partitioning strategy may have introduced systematic differences between the development and test cohorts, and alternative partitioning strategies should be investigated in future studies. Fourth, while the primary framework was rigorously evaluated via five-fold cross-validation, ablation comparisons were conducted on a single representative benchmark partition (Fold 1) due to computational constraints. Nevertheless, the paired evaluation on identical test queries provided sufficient statistical validity to assess inter-model differences. Finally, prospective clinical deployment studies are warranted to evaluate the real-time latency of the system and its practical efficacy in intercepting wrong-patient events in active hospital workflows [4, 5, 11–13].

## 5 Conclusions

This study developed and evaluated an automated patient identity verification framework for cross-modality verification between chest radiographs and CT scout images. While high identification capability was primarily driven by AdaCos-based metric learning, the integration of DADA provided essential structural robustness against severe cross-modality distribution shifts, maintaining high verification and re-identification performance across disparate acquisition geometries and scanner vendors without requiring target-domain identity annotations. The proposed framework operates entirely on routinely acquired diagnostic images without requiring dedicated biometric hardware or extra patient burden. It offers a practical, automated safeguard within PACS and hospital information infrastructures to mitigate patient identification errors, support multimodal data integrity, and enhance clinical safety. Future studies should investigate prospective deployment in real-world healthcare information environments to evaluate its operational efficacy in active hospital workflows.

## Statements and Declarations

## Data Availability

The clinical imaging data used in this study are not publicly available because they contain protected health information and are subject to institutional and ethical restrictions. The data are owned by Yamaguchi University Hospital, Yamaguchi, Japan and cannot be shared publicly.

## Ethics Approval

This study was conducted in accordance with the ethical standards of the institutional research committee and with the 1964 Declaration of Helsinki and its later amendments or comparable ethical standards. All de-identified imaging data previously accumulated for routine clinical care were provided by Yamaguchi University Hospital for this research. Ethics approval for the study protocol of the anonymized dataset was granted by the Institutional Review Board of the University of Osaka Hospital (Approval No. 21064–6).

## Informed Consent

The requirement for informed consent was waived by the Institutional Review Board because of the retrospective nature of the study.

## Declaration of Generative AI and AI-Assisted Technologies in the Writing Process

During the preparation of this manuscript, the authors used Microsoft 365 Copilot (Microsoft Corporation) to assist with language refinement, editing, and manuscript preparation. AI-assisted coding tools were also used during software development associated with this research. All generated outputs were carefully reviewed, validated, and revised by the authors. The authors take full responsibility for the accuracy, integrity, and content of this publication.

## Supporting information

Online Resource 1

## Online Resource 1

### Supplementary Methods, Figures, and Tables

This supplementary material contains the following supporting information for the article:

• Supplementary Methods S1–S3: Detailed mathematical formulation of the deep metric learning and data-augmented domain adaptation (DADA) framework, data augmentation procedures, and training hyperparameters.

• Figure S1: Representative Grad-CAM visualizations demonstrating anatomical regions contributing to patient identity verification.

• Table S1: Representative examples of verification errors and their underlying causes.

## Supplementary Methods

### S1. Deep Metric Learning and Domain Adaptation Framework

The patient identity verification task was formulated as a deep metric learning problem. Given an input image *x*, the feature extractor *f*(⋅) produces a normalized embedding vector:

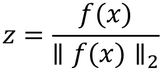

where *z* represents the patient-specific image embedding vector and *f*(*x*) denotes the feature representation extracted from input image *x*.

#### S1.1 AdaCos Loss

Let *z_i_* be a normalized image embedding and *w_j_* be a normalized proxy vector. The cosine similarity is defined as

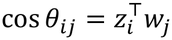

The AdaCos objective function is

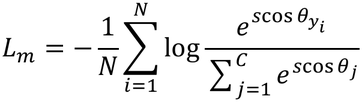

where *s* is the adaptive scale parameter, *y_i_* is the ground-truth identity, and *C* is the number of identity classes.

#### S1.2 Data-Augmented Domain Adaptation

Domain adaptation was performed using data-augmented domain adaptation (DADA) [36]. To reduce the distribution shift between chest radiographs and CT scout images, three domains were aligned during training:

• Source domain (*D_s_*)

• Target domain (*D_t_*)

• Augmented domain (*D_a_*)

The augmented domain was generated through latent-space interpolation between image embeddings and proxy representations.

#### S1.3 Domain Alignment Ablation Models (None and MMD-Based Alignment)

To isolate and validate the individual contribution of the DADA framework (**Table 4**), two comparative domain alignment variants were evaluated under identical training configurations:

##### AdaCos without DADA (None)

The model trained exclusively with the metric learning loss by setting, serving as the baseline to evaluate the intrinsic representation capability of AdaCos without any explicit cross-modality alignment.

##### MMD-Based Alignment

A conventional domain adaptation baseline without data-augmented latent interpolation. Unlike DADA, which aligns source, target, and interpolated augmented domains, this model directly minimizes the discrepancy between the source (CXR) and target (CT scout) proxy-aligned representations using the standard maximum mean discrepancy (MMD):

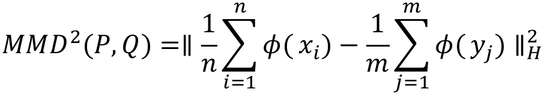

where

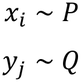

and *φ*(⋅) denotes the feature mapping into the reproducing kernel Hilbert space. The MMD loss weight was set to *λ_da_* = 0.05 to match the regularization strength of the proposed DADA framework.

#### S1.4 Overall Objective Function of the Proposed Framework

For the complete proposed framework, the total optimization objective was:

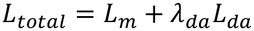

where *L_m_* denotes the AdaCos metric-learning loss and *L_da_* denotes the domain-adaptation loss The weighting coefficient *λ_da_* was fixed at 0.05 according to the original DADA implementation and was not further tuned.

## Supplementary Implementation Details

### S2. Data Augmentation

The training-time data augmentation included random rotation (±5°), random transversal scaling for CT scout images, random perspective distortion for chest radiographs, and top-centered crop-and-pad preprocessing. Before network training, images were resampled to 1.0 × 1.0 mm² and cropped to 448 × 448 pixels.

### S3. Training Configuration

The model was trained for a maximum of 500 epochs, without early stopping, using adaptive sharpness-aware minimization (ASAM) with a stochastic gradient descent (SGD) base optimizer. Gradient accumulation (4 steps) was applied to achieve an effective batch size of 64 from a mini-batch size of 16. A cosine annealing schedule was used to decrease the learning rate from 0.01 to 0.0001. Model selection was based on the minimum validation loss.

#### Summary of Hyperparameters

• **Base Optimizer:** SGD (Momentum = 0.8, Weight Decay = 0.0005)

• **Optimizer Wrapper:** ASAM (Neighborhood size rho = 2.0)

• **Mini-batch Size / Effective Batch Size:** 16 / 64 (Gradient accumulation steps = 4)

• **Learning Rate Schedule:** Cosine annealing (Initial LR = 0.01, Minimum LR = 0.0001)

• **Maximum Epochs:** 500

## Author Contributions

**Conceptualization:** Yasuyuki Ueda

**Methodology:** Yasuyuki Ueda

**Data curation:** Yasuyuki Ueda

**Formal analysis:** Yasuyuki Ueda

**Investigation:** Yasuyuki Ueda

**Writing – original draft preparation:** Yasuyuki Ueda

**Writing – review & editing:** Yasuyuki Ueda

**Supervision:** Takayuki Ishida

All authors read and approved the final manuscript.

## Acknowledgements

The authors thank Masatoshi Yamane and Takuya Uehara of Yamaguchi University Hospital, Yamaguchi, Japan and Yuki Fujimoto of Saiseikai Shiga Hospital, Shiga, Japan for their assistance in organizing the clinical imaging dataset used in this study. The authors also thank Editage (www.editage.com) for English language editing.

## Supplementary Figures

**Figure S1.**
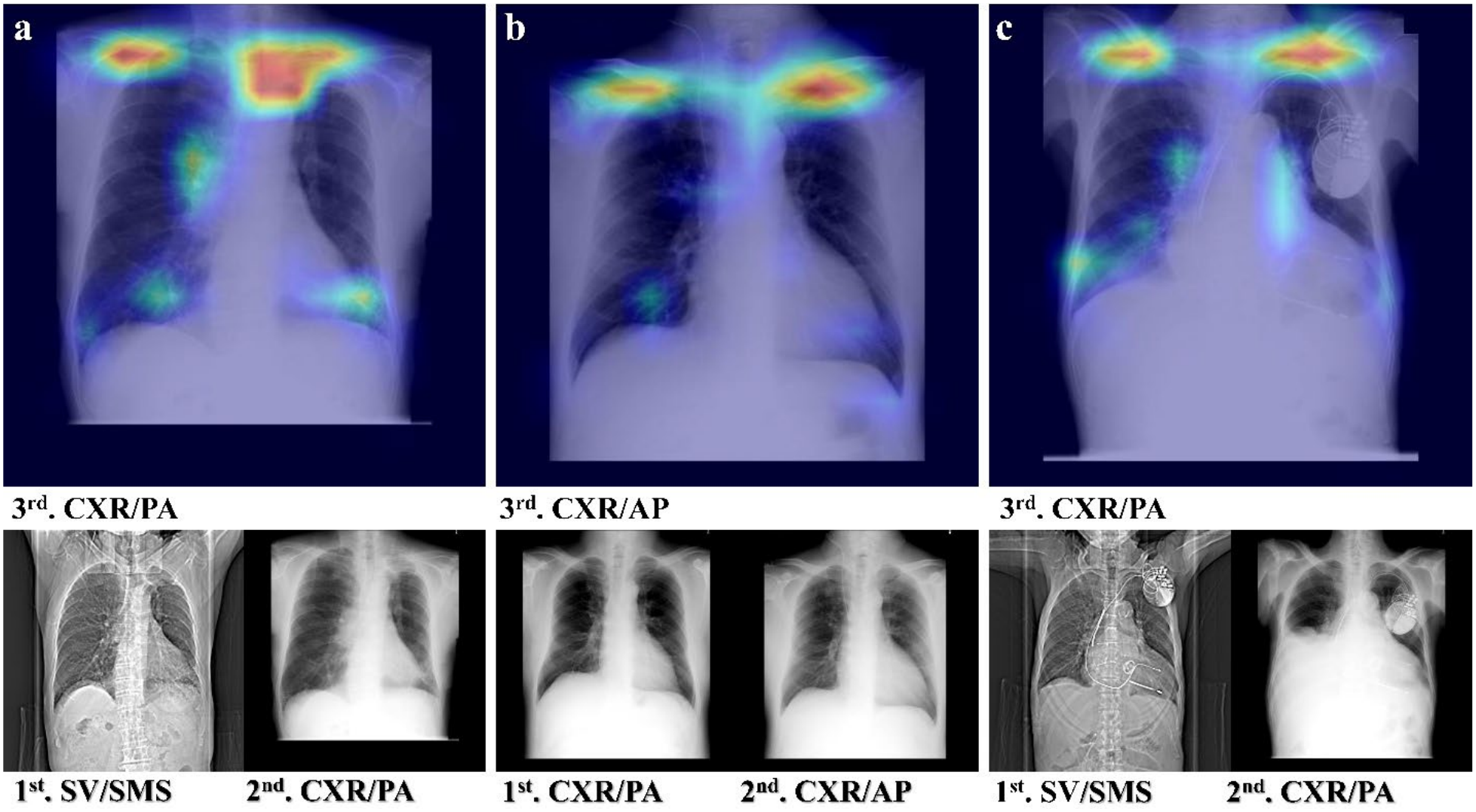
Grad-CAM Analysis. Representative Grad-CAM visualizations of the proposed framework. Grad-CAM heatmaps are superimposed on chest radiographs and CT scout images from three representative patients (**a**–**c**). For each patient, images are shown in chronological order. The first and second images were obtained from the training dataset, whereas the third image was obtained from the validation dataset. The model frequently focused on anatomical structures in the upper thorax, particularly the lung apices and clavicular regions, across imaging modalities. **CXR**: chest radiograph; **SV**: scout view; **SMS**: Siemens Medical Solutions; **GEMS**: GE Medical Systems; **PA**: posteroanterior; **AP**: anteroposterior.

