## Supplementary material for "Automated Patient Identity Verification Framework for Multimodal Medical Imaging Using Deep Metric Learning and Domain Adaptation": Online Resource 1

**Supplementary Methods, Figures, and Tables**

This supplementary material contains the following supporting information for the article:

- Supplementary Methods S1–S3: Detailed mathematical formulation of the deep metric learning and data-augmented domain adaptation (DADA) framework, data augmentation procedures, and training hyperparameters.
- Figure S1: Representative Grad-CAM visualizations demonstrating anatomical regions contributing to patient identity verification.
- Table S1: Representative examples of verification errors and their underlying causes.

### Supplementary Methods

#### S1. Deep Metric Learning and Domain Adaptation Framework

The patient identity verification task was formulated as a deep metric learning problem. Given an input image $x$, the feature extractor $f(\cdot)$ produces a normalized embedding vector:

$$z = \frac{f(x)}{\| f(x) \|_2}$$

where $z$ represents the patient-specific image embedding vector and $f(x)$ denotes the feature representation extracted from input image $x$.

#### S1.1 AdaCos Loss

Let $z_i$ be a normalized image embedding and $w_j$ be a normalized proxy vector. The cosine similarity is defined as

$$\cos\theta_{ij} = z_i^{\top} w_j$$

The AdaCos objective function is

$$L_m = -\frac{1}{N}\sum_{i=1}^{N}\log\frac{e^{s\cos\theta_{y_i}}}{\sum_{j=1}^{C} e^{s\cos\theta_j}}$$

where $s$ is the adaptive scale parameter, $y_i$ is the ground-truth identity, and $C$ is the number of identity classes.

#### S1.2 Data-Augmented Domain Adaptation

Domain adaptation was performed using data-augmented domain adaptation (DADA) [36]. To reduce the distribution shift between chest radiographs and CT scout images, three domains were aligned during training:

- Source domain ($D_s$)
- Target domain ($D_t$)
- Augmented domain ($D_a$)

The augmented domain was generated through latent-space interpolation between image embeddings and proxy representations.

#### S1.3 Domain Alignment Ablation Models (None and MMD-Based Alignment)

To isolate and validate the individual contribution of the DADA framework (**Table 4**), two comparative domain alignment variants were evaluated under identical training configurations:

**AdaCos without DADA (None):** The model trained exclusively with the metric learning loss by setting, serving as the baseline to evaluate the intrinsic representation capability of AdaCos without any explicit cross-modality alignment.

**MMD-Based Alignment:** A conventional domain adaptation baseline without data-augmented latent interpolation. Unlike DADA, which aligns source, target, and interpolated augmented domains, this model directly minimizes the discrepancy between the source (CXR) and target (CT scout) proxy-aligned representations using the standard maximum mean discrepancy (MMD):

$$MMD^2(P,Q) = \| \frac{1}{n}\sum_{i=1}^{n} \phi(x_i) - \frac{1}{m}\sum_{j=1}^{m} \phi(y_j) \|_H^2$$

where

$$x_i \sim P$$

$$y_j \sim Q$$

and $\phi(\cdot)$ denotes the feature mapping into the reproducing kernel Hilbert space. The MMD loss weight was set to $\lambda_{da} = 0.05$ to match the regularization strength of the proposed DADA framework.

#### S1.4 Overall Objective Function of the Proposed Framework

For the complete proposed framework, the total optimization objective was:

$$L_{total} = L_m + \lambda_{da} L_{da}$$

where $L_m$ denotes the AdaCos metric-learning loss and $L_{da}$ denotes the domain-adaptation loss

The weighting coefficient $\lambda_{da}$ was fixed at 0.05 according to the original DADA implementation and was not further tuned.

**Supplementary Implementation Details**

**S2. Data Augmentation**

The training-time data augmentation included random rotation (±5°), random transversal scaling for CT scout images, random perspective distortion for chest radiographs, and top-centered crop-and-pad preprocessing. Before network training, images were resampled to 1.0 × 1.0 $mm^2$ and cropped to 448 × 448 pixels.

**S3. Training Configuration**

The model was trained for a maximum of 500 epochs, without early stopping, using adaptive sharpness-aware minimization (ASAM) with a stochastic gradient descent (SGD) base optimizer. Gradient accumulation (4 steps) was applied to achieve an effective batch size of 64 from a mini-batch size of 16. A cosine annealing schedule was used to decrease the learning rate from 0.01 to 0.0001. Model selection was based on the minimum validation loss.

**Summary of Hyperparameters:**

- **Base Optimizer:** SGD (Momentum = 0.8, Weight Decay = 0.0005)
- **Optimizer Wrapper:** ASAM (Neighborhood size rho = 2.0)
- **Mini-batch Size / Effective Batch Size:** 16 / 64 (Gradient accumulation steps = 4)
- **Learning Rate Schedule:** Cosine annealing (Initial LR = 0.01, Minimum LR = 0.0001)

- **Maximum Epochs:** 500

**Supplementary Figures**

***Grad-CAM Analysis***

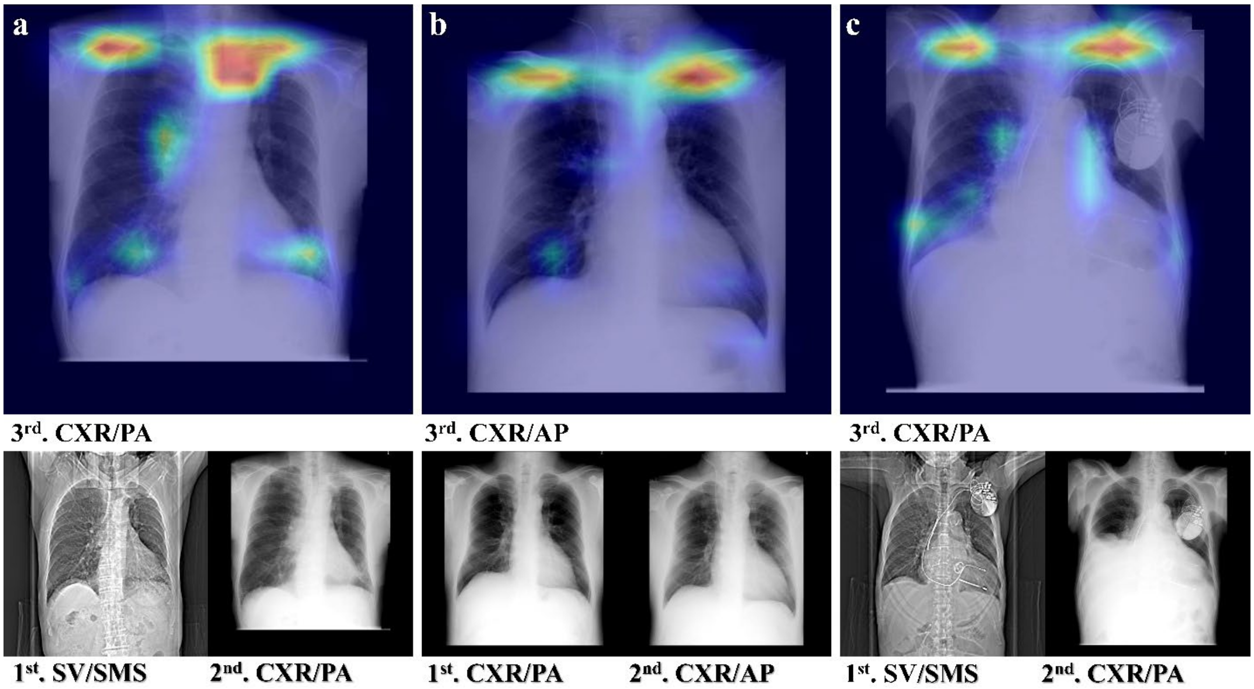


**Figure S1.** Representative Grad-CAM visualizations of the proposed framework. Grad-CAM heatmaps are superimposed on chest radiographs and CT scout images from three representative patients (**a**–**c**). For each patient, images are shown in chronological order. The first and second images were obtained from the training dataset, whereas the third image was obtained from the validation dataset. The model frequently focused on anatomical structures in the upper thorax, particularly the lung apices and clavicular regions, across imaging modalities. **CXR**: chest radiograph; **SV**: scout view; **SMS**: Siemens Medical Solutions; **GEMS**: GE Medical Systems; **PA**: posteroanterior; **AP**: anteroposterior.
